# Spatiotemporal analyses illuminate the competitive advantage of a SARS-CoV-2 variant of concern over a variant of interest

**DOI:** 10.1101/2021.09.14.21262977

**Authors:** Alexis Russell, Collin O’Connor, Erica Lasek-Nesselquist, Jonathan Plitnick, John P. Kelly, Daryl M. Lamson, Kirsten St. George

**Author notes:** These authors contributed equally to this work (Alexis Russell, Collin O’Connor, Erica Lasek-Nesselquist).

## Abstract

The emergence of novel SARS-CoV-2 variants in late 2020 and early 2021 raised alarm worldwide and prompted reassessment of the management, surveillance, and projected future of COVID-19. Mutations that confer competitive advantages by increasing transmissibility or immune evasion have been associated with the localized dominance of single variants. Thus, elucidating the evolutionary and epidemiological dynamics among novel variants is essential for understanding the trajectory of the COVID-19 pandemic. Here we show the interplay between B.1.1.7 (Alpha) and B.1.526 (Iota) in New York (NY) from December 2020 to April 2021 through phylogeographic analyses, space-time scan statistics, and cartographic visualization. Our results indicate that B.1.526 likely evolved in the Bronx in late 2020, providing opportunity for an initial foothold in the heavily interconnected New York City (NYC) region, as evidenced by numerous exportations to surrounding locations. In contrast, B.1.1.7 became dominant in regions of upstate NY where B.1.526 had limited presence, suggesting that B.1.1.7 was able to spread more efficiently in the absence of B.1.526. Clusters discovered from the spatial-time scan analysis supported the role of competition between B.1.526 and B.1.1.7 in NYC in March 2021 and the outsized presence of B.1.1.7 in upstate NY in April 2021. Although B.1.526 likely delayed the rise of B.1.1.7 in NYC, B.1.1.7 became the dominant variant in the Metro region by the end of the study period. These results reveal the advantages endemicity may grant to a variant (founder effect), despite the higher fitness of an introduced lineage. Our research highlights the dynamics of inter-variant competition at a time when B.1.617.2 (Delta) is overtaking B.1.1.7 as the dominant lineage worldwide. We believe our combined spatiotemporal methodologies can disentangle the complexities of shifting SARS-CoV-2 variant landscapes at a time when the evolution of variants with additional fitness advantages is impending.

## Introduction

The emergence of a novel SARS-CoV-2 variant B.1.1.7 (Alpha) in the United Kingdom (UK) in late 2020 raised alarm worldwide and prompted major reassessment of the management, surveillance, and projected future of COVID-19 (1,2). Evidence of increased transmissibility and potential immune evasion prompted the World Health Organization to designate B.1.1.7 a variant of concern (VOC) in December 2020 (3–6). Increased transmissibility of B.1.1.7 is likely due to several mutations, including N501Y which confers antibody evasion (7,8) and increases spike protein binding to the host cell (9), and del60-70 which enhances infectivity (10). The emergence of B.1.1.7 and additional novel SARS-CoV-2 variants with competitive advantages have resulted in localized dominance of single variants (11) and raised concern for increases in COVID-19 incidence (12).

Around the time B.1.1.7 emerged and became dominant in the UK, novel variant B.1.526 (Iota) arose from within New York State (NY) (13,14). The proportion of B.1.526 quickly increased in New York City (NYC) and led to a noticeable shift in lineage distribution during early 2021 (13,14). The World Health Organization designated B.1.526 as a variant of interest (VOI) due to its increase in prevalence coupled with mutations associated with immune evasion (15). Spike mutation E484K, which is present in approximately 45% of B.1.526 genomes sequenced in NY (GISAID.org) and is shared with the B.1.315 (Beta) and P.1 (Gamma) VOCs, is associated with reduced neutralization by monoclonal antibodies (16). Further, 29% of B.1.526 sequenced samples in NY possess the S477N spike mutation (GISAID.org), which likely evolved twice independently within the lineage (13). S477N is hypothesized to confer increased transmissibility due to its location at the ACE2 binding receptor in spike, which is required for viral entry into the host cell. (17). Despite these concerns, an epidemiological assessment of B.1.526 in NYC from January through April 2021 found that the lineage did not cause more severe disease and was not associated with increased risk of reinfection or vaccine breakthrough (18).

Genomic surveillance of COVID-19 is a crucial tool to monitor and assess the physiological and epidemiological characteristics of SARS-CoV-2 variants as they emerge. In order to track the spread and impact of novel variants in NY, the New York State Department of Health (NYSDOH) significantly expanded its genomic surveillance program in December 2020, with the aim of sequencing a more representative subset of COVID-19 cases across the state. A robust genomic surveillance system allows for the assessment of changes in variant distribution over precise temporal and spatial scales. The co-circulation of B.1.526 and B.1.1.7 within NY in late 2020 and early 2021 offers a unique opportunity to compare variant transmission dynamics from a phylogeographic and spatial-epidemiological context. While phylogeographic analyses are routinely employed to infer the timing and source of SARS-CoV-2 introductions around the world (19–22), spatial statistics offer a complementary approach for examining COVID-19 incidence and the spatial spread of SARS-CoV-2 variants. Spatial statistics, including scan statistics for detecting clusters, are often employed in an epidemiological context to assess whether disease incidence is higher in one area than others (23). The discrete Poisson space-time scan statistic is a spatial statistical method that has been adopted to search for geographic areas with high incidence of COVID-19 (24,25). Similarly, the multinomial scan statistic is commonly used for the purposes of detecting geographic areas with unusually high distributions of specific disease or event types. The multinomial scan statistic has been used to clarify categorical distributions in a variety of contexts, including the distribution of meningitis types (26), multiresistance phenotypes of Staphylococcus aureus (27), and methods of suicide (28). To the best of our knowledge, the use of multinomial scan statistic to quantitatively assess the categorical distribution of SARS-CoV-2 variants among total COVID-19 cases has not been reported. This study employs spatial scan statistics paired with phylogeographic analyses to describe the shifting SARS-CoV-2 variant landscape in NY from December 2020 to April 2021, particularly the inter-play between B.1.526 and B.1.1.7. We identify the sources of multiple introductions of B.1.1.7 and B.1.526 in NY and track their spatiotemporal spread relative to oneanother. As evidenced by the rapid dominance of the highly transmissible VOC Delta in many locations around the world in the summer of 2021 (GISAID.org), inter-variant competition will likely play a major role in the future public health response and epidemiology of COVID-19. Our findings elucidate the dynamics of competing SARS-CoV-2 variants at a time when the future variant landscape remains uncertain.

## Results

A total of 8,517 SARS-CoV-2 specimens sequenced by Wadsworth with collection dates between December 1, 2020 and April 30, 2021 were included in the study. Sequenced specimens equated to 0.6% of new COVID-19 cases in the study period. Among the included specimens, B.1.1.7 and B.1.526 constituted 1,107 (13%) and 904 (10.6%) of the samples, respectively. Formerly defined parent lineage B.1.526 and sublineages B.1.526.1, B.1.526.2, and B.1.526.3 constituted 98, 388, 415, and 2 samples, respectively. The average patient age for all samples was 46.5 years, which decreased from 49.6 years in December to 39.5 years in April. Patient age and week of sample collection exhibited a negative association during the study (*β*=0.765, t= 16.31, p<0.0001). The mean age of sampled patients also differed significantly between variants (df=2, F=81.39, p<0.0001), where mean ages for B.1.1.7, B.1.526, and all other lineages were 39.3, 44.6, and 48.5 years, respectively.

The earliest B.1.1.7 samples sequenced by Wadsworth were collected on December 24, 2020 from a resident of Manhattan (Metro region) and an individual in Saratoga County (Capital region). B.1.1.7 remained relatively rare among all samples through the end of January. The Metro and Capital regions experienced the earliest increases in B.1.1.7, although the proportion of B.1.1.7 did not exceed 15% through February. The proportion of B.1.1.7 increased in March across all regions, most notably in the Western region where it constituted around 75% of all samples by the end of March and continued to rise through April. The Metro region experienced the most gradual increase in B.1.1.7, with the proportion not exceeding 40% until the end of April.

The earliest B.1.526 sample sequenced by Wadsworth was collected on December 9, 2020 from a patient in the Bronx (Metro region). The proportion of B.1.526 increased in the Metro region throughout December, reaching 10% of total samples by the end of the month. The proportion of B.1.526 in the Metro region approached 40% by the end of January, peaked at ∼60% in mid-February to early March, and then plateaued around 50% through April. In contrast, B.1.526 was not consistently detected in the other regions until February, with increases after February in the Capital and Central regions, but the proportion generally remained under 40%. The Western region saw a minimal increase in B.1.526 after February, with the proportion remaining under 15% through April. The combined proportion of all lineages other than B.1.1.7 and B.1.526 dropped below 20% in all NY regions by the end of April.

Maps of interpolated proportion of B.1.1.7 relative to all other lineages by ZCTA (Figure 1A) show a general trend of spread through the southern portion of NY in January, statewide distribution by February, diffuse increase in proportion in March, and a sustained high proportion throughout the state in April, with strong dominance in the Western region. In contrast, maps of interpolated proportion of B.1.526 shows more constricted initial spread focused around NYC and surrounding areas in January, with statewide distribution not achieved until March, and a moderate proportion sustained mostly within the Metro region, without notable increase in proportion from March to April (Figure 1B).

**Figure 1:**
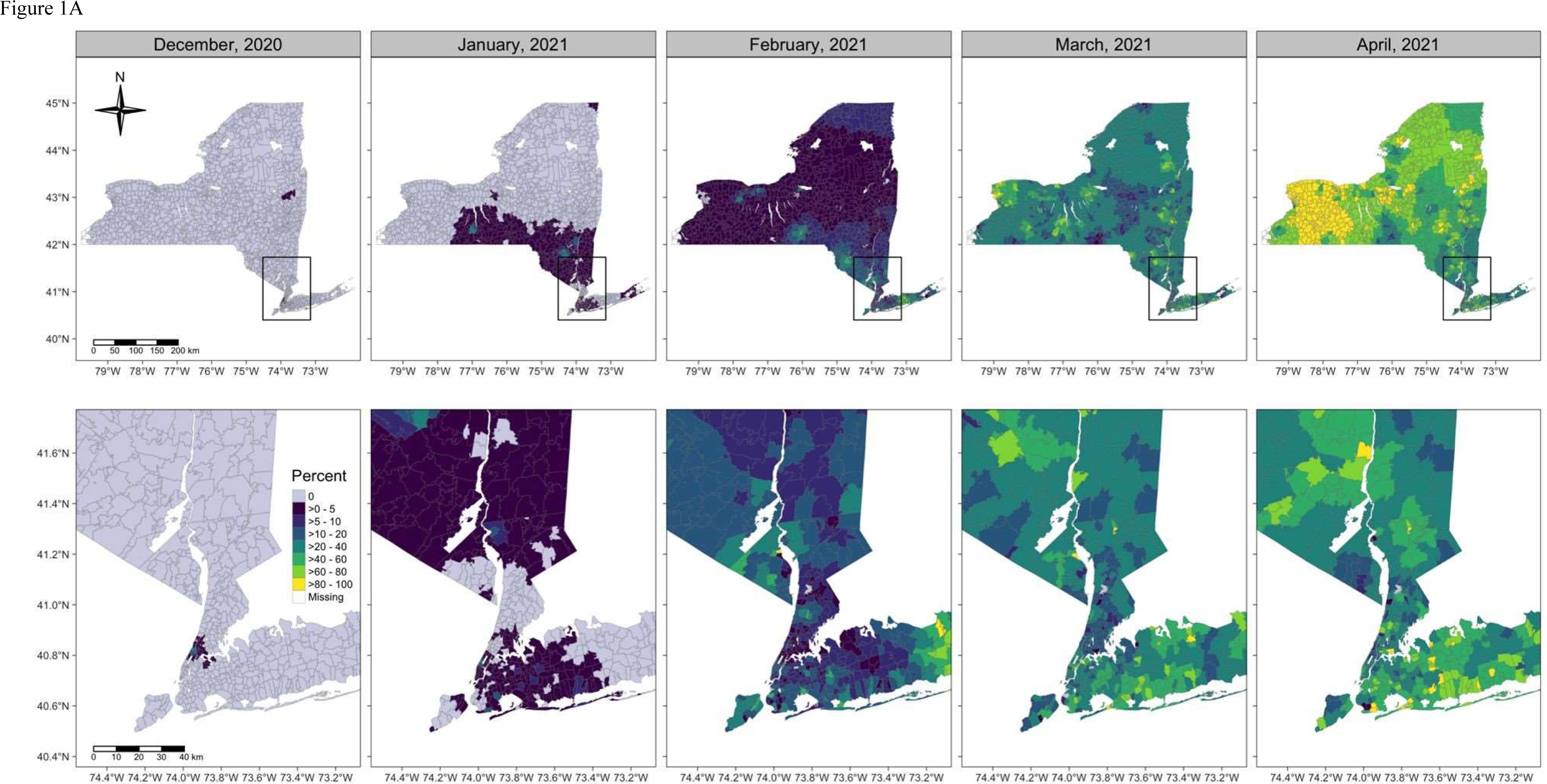
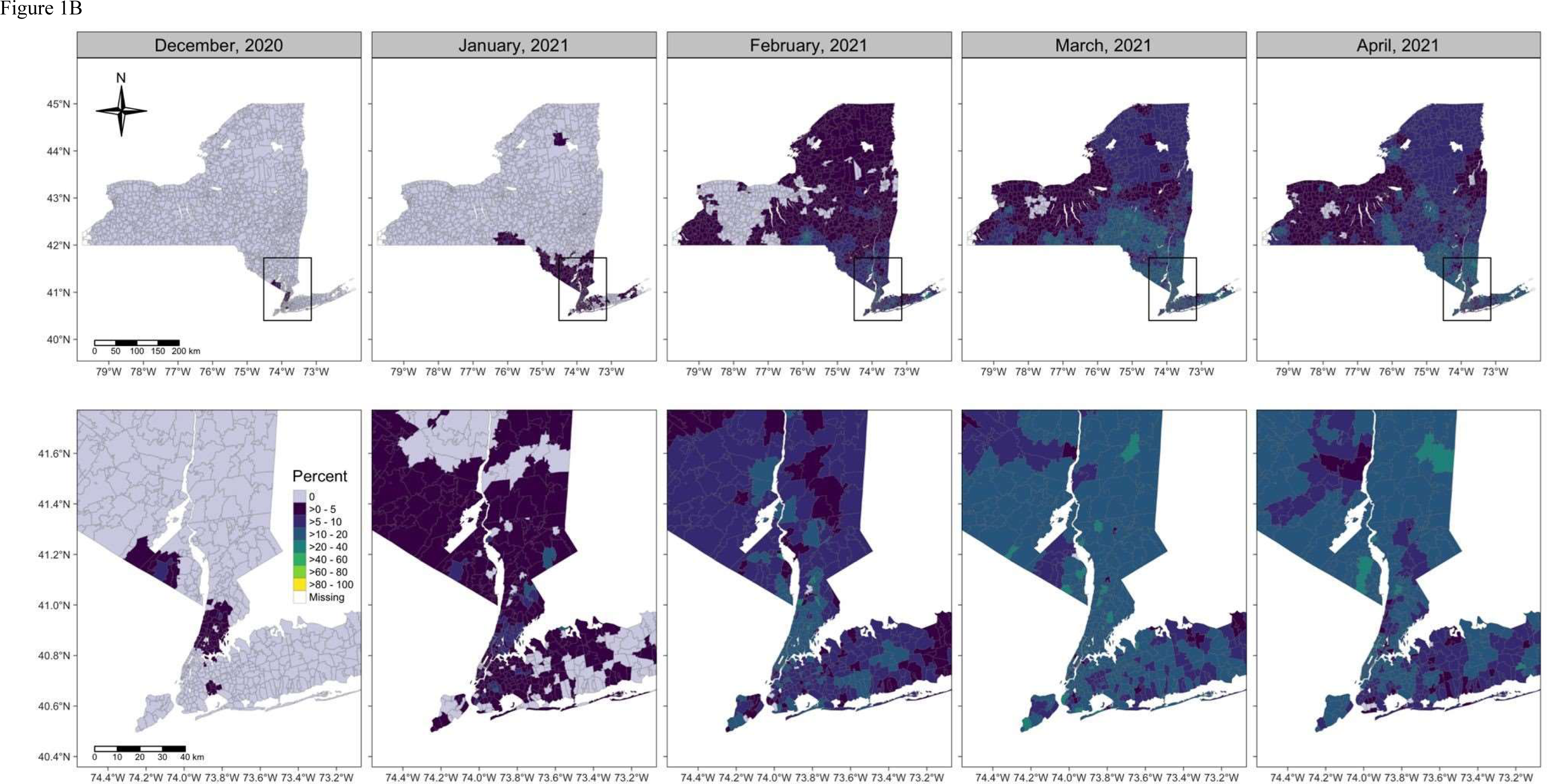
Inverse distance weighted interpolations of A) percent of B.1.1.7 and B) percent of B.1.526 relative to all other lineages by ZIP code tabulation area.

Maps of geographic mean centers of estimated B.1.1.7 and B.1.526 cases (Figure 2) show that shifts in the SARS-CoV-2 variant landscape had an impact on the spatial distribution of overall cases of COVID-19. The mean center of B.1.526 cases initially occurred near the NYC area, then gradually moved slightly northwest as B.1.526 expanded modestly into upstate regions. The mean center of B.1.1.7 cases was also in the NYC area at the beginning of the study period, then moved northwest during March and April to a much greater degree than for B.1.526 cases, indicating more substantial expansion of B.1.1.7 than B.1.526 outside of NYC. The mean centers of all COVID-19 cases showed a southeastern trajectory during December through March, but then exhibited a large northwestern shift in April, suggesting that the spread of B.1.1.7 in Upstate NY, especially within the Western region, resulted in a spatial shift of overall COVID-19 cases.

**Figure 2:**
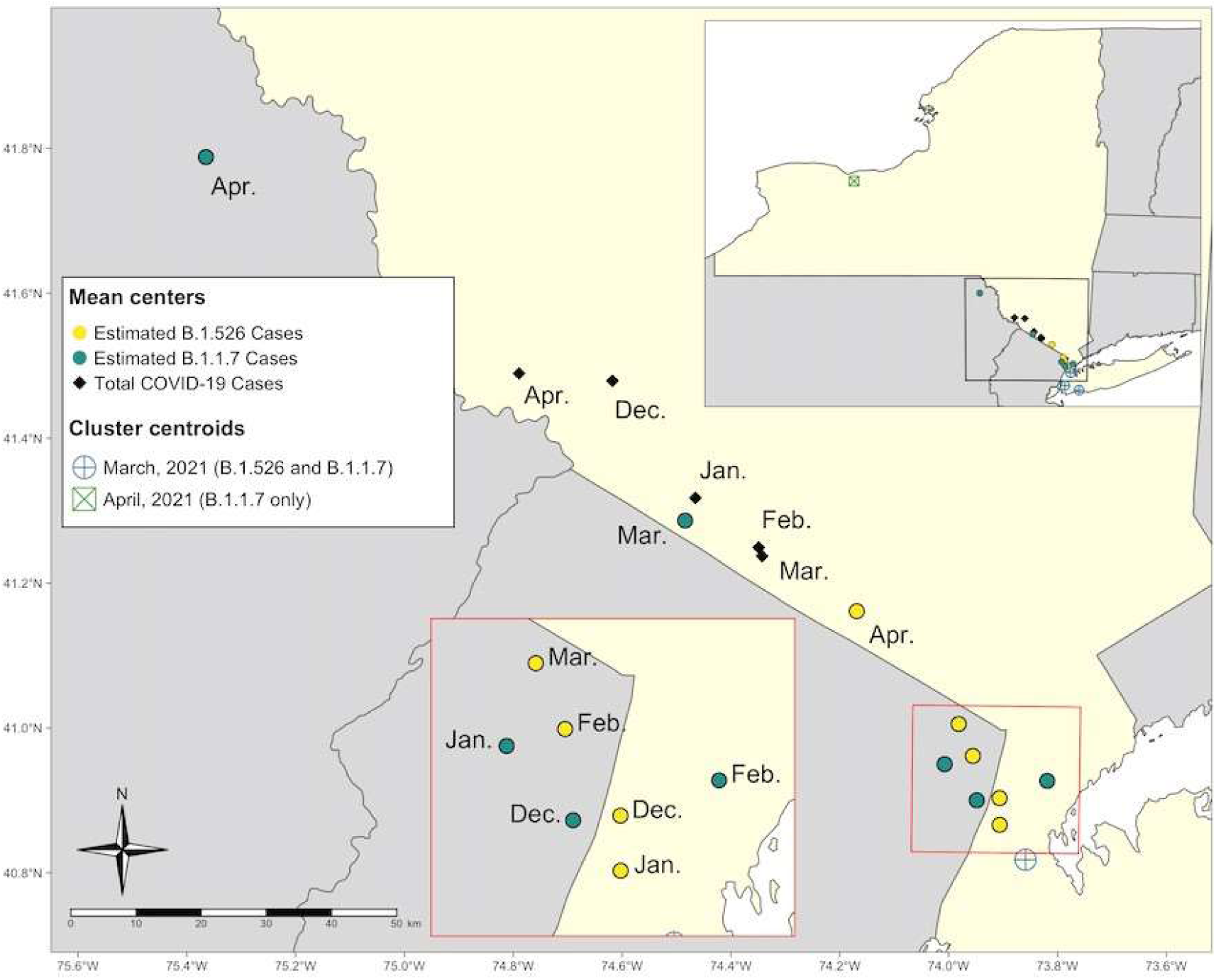
Geographically weighted mean centers of total COVID-19 cases and estimated COVID-19 cases attributable to B.1.526 and B.1.1.7, December 2020 through April 2021. Cluster centroids refer to the results of the multinomial space-time scan analysis (Figure 3).

Retrospective multinomial space-time scan analysis indicated six statistically significant clusters with elevated relative risk (RR) of COVID-19 attributable to specific variants (Figure 3 and Table 1S). Two clusters of elevated RR of “Other” lineages were found in December, 2020 in the Metro/Capital regions as well as Long Island, reflecting the nearly non-existent risk for B.1.1.7 and B.1.526. Of note, these clusters are likely to be limited in spatial extent due to setting the maximum cluster size to 10% of the population-at-risk, as the presence of B.1.1.7 and B.1.526 were also nearly non-existent statewide in December (Figure 1). Three clusters of elevated RR of multiple combinations of B.1.1.7, B.1.526 parent lineage, B.1.526.1, and B.1.526.2 were found in March, 2021 in the NYC/Long Island region. The sixth cluster exhibited an elevated RR of greater than 7.0 for B.1.1.7, with a radius of 114.38 km centered in the Finger Lakes area (Western and Central regions) during April. Additionally, the presence of clusters of B.1.1.7 and B.1.526 in March and April coincide with a general statewide decrease in incidence of COVID-19 (Figure 3).

**Figure 3:**
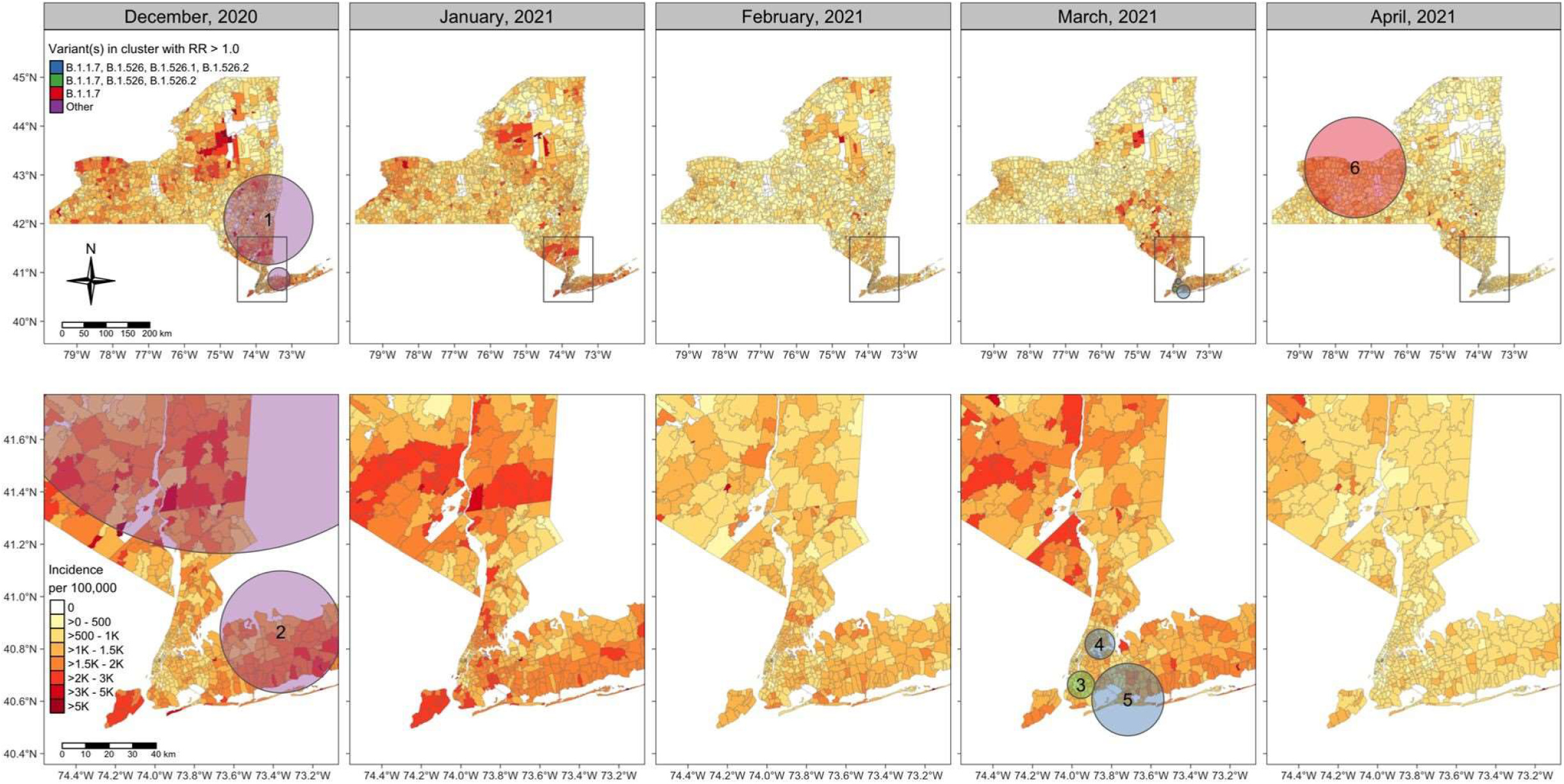
SARS-CoV-2 variant clusters identified from retrospective multinomial space-time scan analysis and COVID-19 incidence by ZIP code tabulation area, December 2020 through April 2021.

The final B.1.526 dataset for phylogenetic reconstruction contained 980 genomes from all regions of NY and various domestic locations (Bronx: 222, Hudson Valley: 128, Brooklyn: 39, Long Island: 78, Manhattan: 49, Queens: 81, Staten Island: 12, Upstate NY: 81, Domestic: 290). The final B.1.1.7 dataset contained 1,195 genomes from the NYC region (181), Finger Lakes (239), Hudson Valley (78), Long Island (130), Western NY and the Southern Tier (Southwestern NY: 56), Capital District, Mohawk Valley, Central NY and the North Country (Northern NY: 149), as well as other states (Domestic: 362). Results from the phylogeographic analysis indicated that B.1.526 emerged within the Bronx near the end of 2020, and that this location was the major source of spread to other regions of NY and US (Domestic) in the ensuing months (Figure 4A). Although sampling biases could have influenced the number of introductions assigned to the Bronx, the Domestic category had greater representation in the dataset but led to substantially fewer introductions (Table S2). Specifically, Domestic genomes represented 29.5% of the dataset but this location was responsible for only 6.7% of all B.1.526 introductions, while the Bronx represented 22.7% of the dataset and led to 63.8% of all introductions (Table S2). Excluding the Bronx, B.1.526 transmission between boroughs and from these boroughs to other NY or Domestic locations was relatively infrequent. Further, introductions were mostly uni-directional, with only 9% of the B.1.526 cases sequenced from the Bronx due to re-introductions. This indicates the spread of B.1.526 in the Bronx was overwhelmingly attributable to transmission within the Bronx itself and contrasts the trends for other NY and Domestic locations, which showed more limited sustained transmission of B.1.526 as a function of the ratio of number of introductions to sample size.

**Figure 4:**
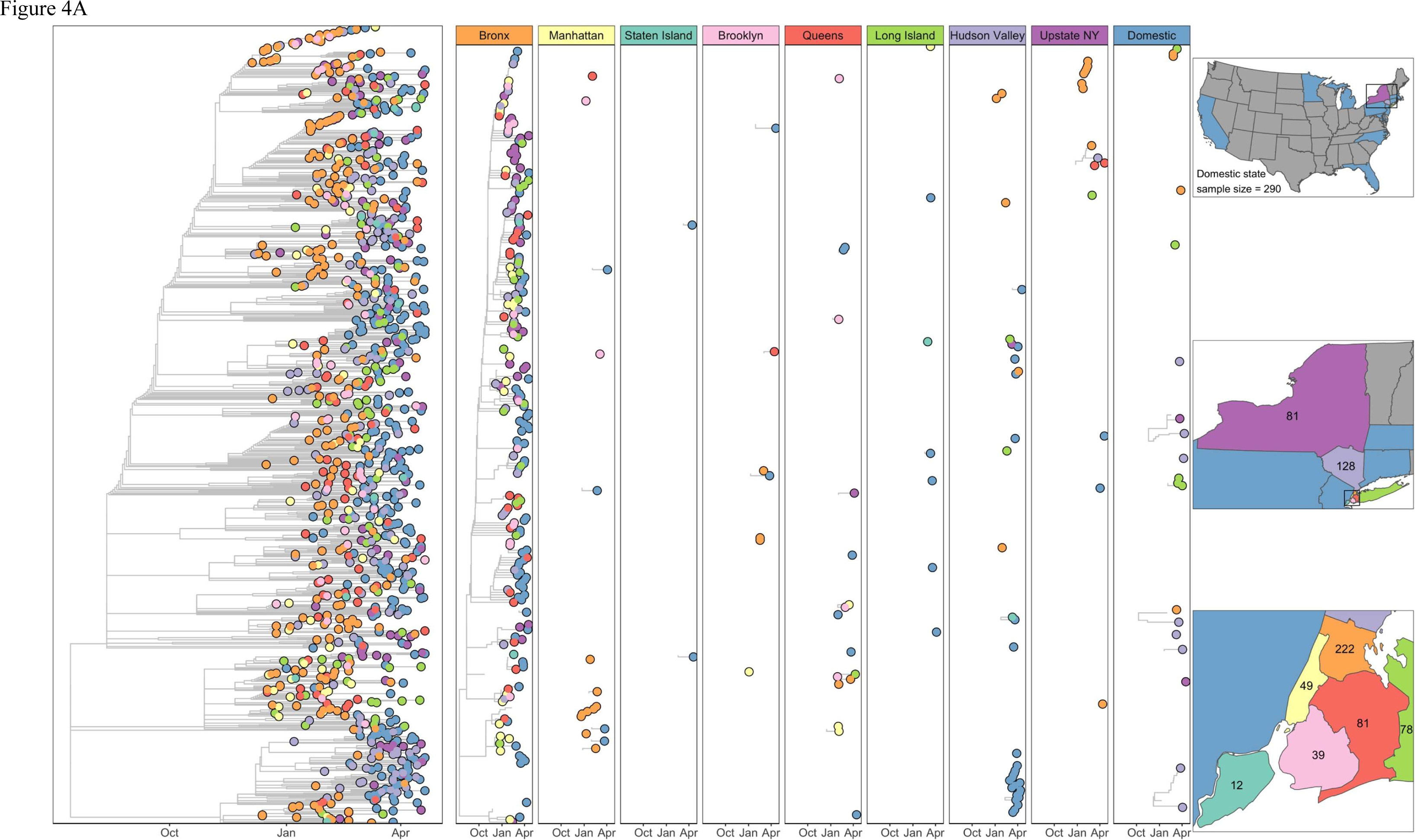
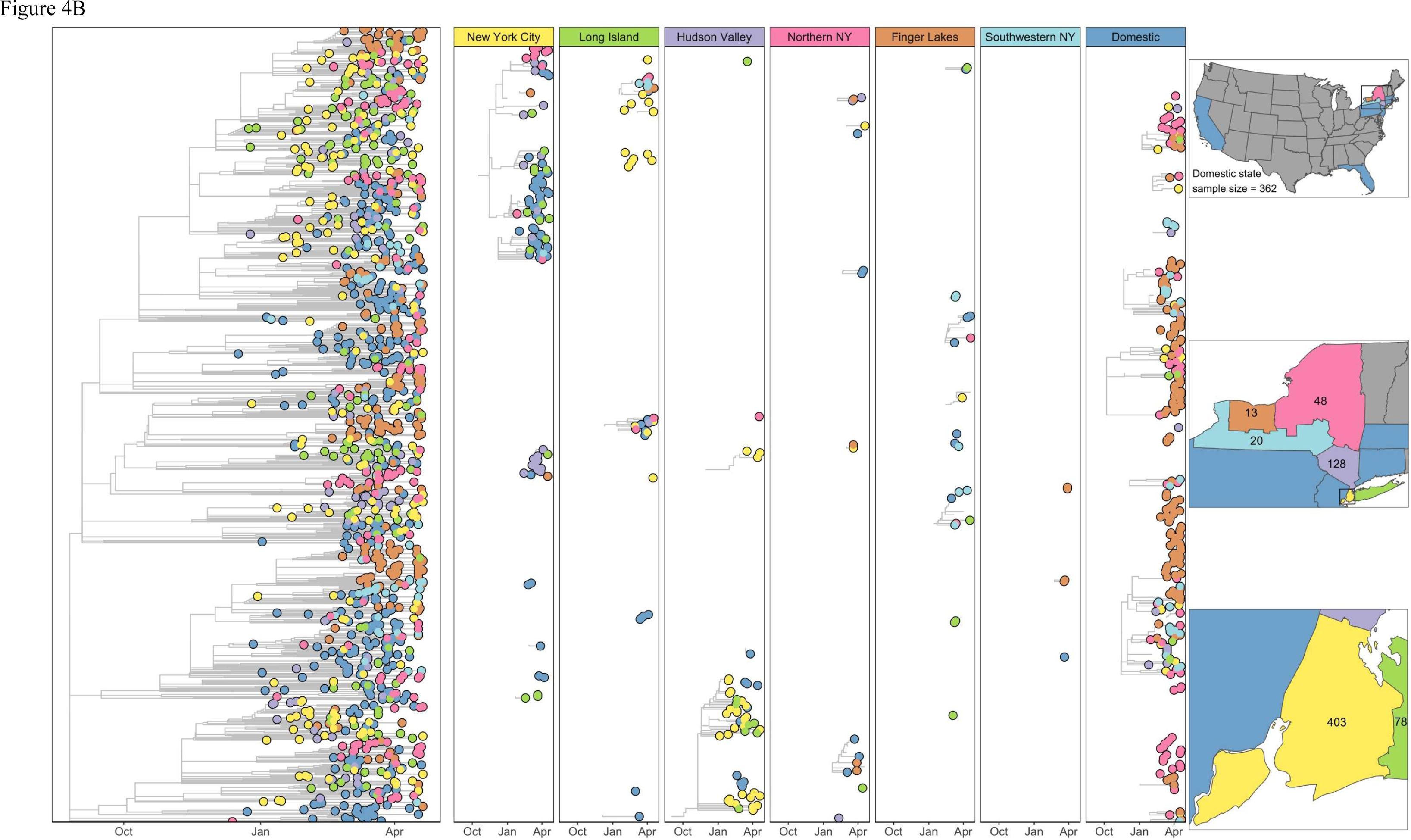
A)Time-calibrated phylogeny of SARS-CoV-2 variant B.1.526. Left panel represents a maximum likelihood phylogeny of 908 genomes from New York and other states generated in IQ-tree with timescale inferred by TreeTime and ancestral state reconstruction performed in BEAST. Faceted panels indicate the source of B.1.526 introductions into different region of New York or other states (Domestic). Only introductions supported by an ancestral state probability of >= 0.7 are shown. Right panel shows locations sampled and sample sizes. B) Time-calibrated phylogeny of SARS-CoV-2 variant B.1.1.7. Left panel represents a maximum likelihood phylogeny of 1,195 genomes from New York and other states generated in IQ-Tree with timescale inferred by TreeTime and ancestral state reconstruction performed in BEAST. The tree was rooted with a P.1 genomes (not shown). Faceted panels indicate the source of B.1.1.7 introductions into different regions of New York and to other states (Domestic). Only introductions supported by an ancestral 414 state probability of >= 0.7 are shown. Right panel shows locations sampled and sample sizes.

Multiple domestic introductions contributed to the initial presence of B.1.1.7 in NY (29), with transmission occurring most frequently to the Finger Lakes and Northern NY (Figure 4B). The Finger Lakes and Northern NY were well-represented in the dataset (20% and 12% of the data, respectively) but contributed substantially less to the distribution of B.1.1.7 (accounting for 7% and 6% of the total number of introductions, respectively) than Domestic sites, which represented 20% of the data and were responsible for the majority of introductions (∼39%, Table S3). Exchange between NYC and Long Island and NYC and the Hudson Valley was also frequent, but transmission from these regions to Northern NY, Southwestern NY, and the Finger Lakes was substantially more limited (Figure 4B, Table 3), possibly reflecting the effects of interconnectivity in a metropolitan area dominated by commuter activity. The Finger Lakes showed the lowest proportion of sequenced cases due to introductions but the largest sample size in NY, suggesting more sustained transmission of B.1.1.7 within this region. The topologies of the B.1.526 and B.1.1.7 phylogenetic trees highlight differences in transmission patterns as well. Introductions of B.1.526 often represent single events not associated with additional transmission (except for some introductions from the Bronx), which led to B.1.526 genomes from various locations being distributed broadly throughout the tree. In contrast, introductions of B.1.1.7 often led to large regional clusters, such as those in the Finger Lakes from Domestic sources, producing a patchier distribution of genomes from the same region.

## Discussion

The repeated emergence of novel variants of SARS-CoV-2 has largely defined the COVID-19 pandemic response in 2021. As vaccination rates, prior exposure levels, and behavioral public health measures continuously change, so too does the strength of selection (30). Given that selective pressures likely vary across regions, including between and within states, it follows that the emergence and spread of SARS-CoV-2 variants are also regionally dynamic. We combine spatial statistical, phylogeographic, and cartographic visualization techniques to examine the spatiotemporal dynamics of the VOC B.1.1.7 (Alpha) and the VOI B.1.526 (Iota) within NY from December 2020 through April 2021. Our study period captures a time of substantial fluctuations in statewide COVID-19 incidence, with a major peak in late December and early January followed by decline through February, and then a smaller peak in late March and early April followed by a decline through April. The peak incidence in late 2020 and early 2021 represents the second major wave of COVID-19 cases in NYC, and the first major wave for much of the rest of the state. Our study period also spans the launch of NY’s vaccination campaign in January 2021 and subsequent rise in immunization rates.

The concurrent spread of B.1.1.7 and B.1.526 offers a unique opportunity to compare the dynamics of two competing variants of SARS-CoV-2 within a population. Shortly after its appearance in the Bronx in late 2020, B.1.526 quickly became the most common lineage in NYC and the surrounding region by mid-February. The rapid dominance of B.1.526 within NYC is corroborated by time-calibrated phylogeny (Figure 4A), which depicts widespread initial transmission within the Bronx, periodic introductions to neighboring boroughs, and later introductions to the greater Metro region and other states, with few introductions leading to sustained transmission outside the Bronx. The spread of B.1.526 appears to have been spatially limited by the repeated introduction and apparent transmission advantage of B.1.1.7 outside of NYC. Regions of NY where B.1.526 had not yet established experienced rapid dominance of B.1.1.7 during March and April. This trend is most clearly seen in the near complete displacement of all other lineages by B.1.1.7 in Western NY (Supplemental Figure 1A), resulting in a large cluster of elevated RR for B.1.1.7 cases in the Finger Lakes region during April (Figure 3). Phylogeographic analysis provides support for the sustained transmission of B.1.1.7 in the Finger Lakes, evidenced by the large regional clusters resolved in the tree. The multinomial spatial scan detected three unique clusters in March 2021, all with increased RR for B.1.1.7 and B.1.526. The values for RR within each NYC cluster details a distinct pattern: clusters centered within the Bronx, Brooklyn, and Manhattan had higher RR for B.1.526, while the cluster centered in east Queens and Long Island had a higher RR for B.1.1.7 (Table S1). Over the months following B.1.526’s initial advantage in NYC, B.1.1.7 trends towards become the majority variant in the Metro region. Despite the presence of both B.1.1.7 and B.1.526 in the Metro region in the early months of 2021, phylogeographic analyses indicated that the parental lineage of B.1.526 was circulating in the Bronx by August 2020, with more frequent transmission to Manhattan than Long Island. While B.1.1.7 was circulating in Long Island by mid-October 2020, almost all B.1.526 introductions to this region occurred in 2021. Thus, there are important distinctions between the spread of B.1.1.7 and B.1.526 even within a geographically proximal area. The delayed dominance of B.1.1.7 in the Bronx compared to Long Island and Queens suggests that B.1.526 was more difficult to displace than other lineages circulating in these areas. The number of B.1.1.7 introductions into NYC was greater than into the Finger Lakes, but sustained transmission was less prevalent, providing further evidence that widespread circulation of B.1.526 in NYC stunted the initial spread of B.1.1.7 in this area. Maps of the geographic mean centers of the estimated number of COVID-19 cases attributable to each variant capture the rapid spread of B.1.1.7 out of NYC, as well as the relative inability of B.1.526 to claim a foothold outside of the Metro region. The northwesterly shift in the trajectory of overall COVID-19 cases in April indicates that the expansion of B.1.1.7, in particular clustering in Western NY, had a measurable influence on the spatial spread of COVID-19 cases overall.

There are several limitations of our study which primarily reflect the inherent limitations of our genomic surveillance program. A degree of selection bias exists within our dataset given that specimens were screened by cycle threshold value and were submitted by a selected group of clinical and commercial labs that cannot perfectly represent all COVID-19 cases in NY. We were unable to assess the demographic and clinical representativeness of our dataset because these data were not available to us for many specimens. Additionally, the number of specimens sequenced varied over the space and time of the study period, which created small sample sizes within many ZCTA-months. This limitation extended to the multinomial scan statistic, which was run with estimated values for COVID-19 cases attributable to B.1.1.7 and B.1.526, giving all ZCTAs with samples equal weight. However, the spatial scan assesses data according to their proximity to each other. In this context, ZCTAs are analyzed together rather than individually, which has the potential to reduce bias. Another consequence of our limited sampling was that our data exhibited zero samples from many ZCTAs for each month. We addressed this by using IDW interpolation of the proportion of B.1.1.7 and B.1.526 sequenced samples at the ZCTA-month level to visualize general patterns of variant proportions over geography.

Phylogeographic analyses were hampered by similar limitations: uneven sampling among regions and the lack of global representation in our datasets could lead to incorrect trait assignments. Smaller sample sizes for some regions might have caused an underestimation of their contributions to variant transmission in NY while larger sample sizes might have inflated the number of introductions assigned. However, we believe the results presented here largely capture the transmission dynamics of B.1.526 and B.1.1.7 within NY, as smaller sample sizes were consistent with lower incidence, while larger sample sizes did not always correspond to regions with outsized contributions to the spread of either variant.

Our results are in general agreement with those of Petrone et al. 2021, who showed that B.1.1.7 had a higher reproductive value and was able to spread faster than B.1.526 in Connecticut, where both variants were introduced a similar number of times. The reproductive value for B.1.1.7 in NYC was also higher than B.1.526 but the data were noisier, which supports our conclusions that the clear advantage B.1.1.7 showed in Western NY and the Finger Lakes was obfuscated in NYC by the initial dominating presence of B.1.526.

Our phylogeographic and spatiotemporal analyses offer a methodology for evaluating the relative transmissibility and competitive advantages of co-circulating SARS-CoV-2 variants. We demonstrate that the emergence of VOI B.1.526 slowed the rise of VOC B.1.1.7 within NYC, while B.1.1.7 simultaneously became the majority variant in parts of NY devoid of B.1.526. In this way, our study describes important dynamic interactions between variants with unequal transmissibility and is potentially generalizable to interactions between any known variant and the highly transmissible B.1.617.2 (Delta) variant and other variants to come. As B.1.617.2 continues to change the variant landscape on local and global levels, understanding its dynamic interactions with other variants is increasingly important in the management of COVID-19.

## Methods

### Sample acquisition and RNA extraction

The NYSDOH Wadsworth Center coordinated with over 30 clinical laboratories throughout NY who routinely submitted respiratory swabs positive for SARS-CoV-2 for whole genome sequencing. Specimens were required to have a real-time cycle threshold value less than 30. Nucleic acid extraction was performed on a Roche MagNAPure 96 with the Viral NA Small Volume Kit (Roche, Indianapolis, IN) with 100μL sample input and 100μL eluate.

### Sequencing and bioinformatics processing

Extracted RNA was processed for whole genome sequencing with a modified ARTIC protocol (artic.network/ncov-2019) in the Applied Genomics Technology Core at the Wadsworth Center as previously described (29); supplementary information). Illumina libraries were processed with ARTIC nextflow pipelines (github.com/connorlab/ncov2019/articnf/tree/illumine, last updated April 2020) as previously described (Alpert et al., 2021; supplementary information)

### Sample inclusion criteria

Specimens with collection dates between December 1, 2020 and April 30, 2021 were included. Specimens that were sequenced as a result of pre-screening for specific mutations or clinical/epidemiological criteria were removed from the analysis. In the case of duplicate specimens from the same patient, the earliest collected specimen was included, and all other specimens excluded from the analysis. Only specimens with ZIP code of patient address available were included.

### COVID incidence calculation

Monthly COVID case counts by ZIP code were obtained from https://gibhub.com/nychealth/coronavirus-data for NYC, and from the NYSDOH Communicable Disease Electronic Surveillance System for the remainder of NY. Reports with case status of ‘confirmed’ or ‘probable’ were included in the case count. Cases were assigned month based on date of diagnosis. All ZIP code data was converted to ZIP code tabulation area (ZCTA). Incidence was calculated using ZCTA-level population data from the 2019 1-year American Community Survey estimates.

### Retrospective multinomial space-time scan statistic

We utilize the retrospective multinomial space-time scan statistic in SaTScan version 9.6, using the non-ordinal method (26,31). Estimated SARS-CoV-2 variant data used in the multinomial scan statistic were calculated for each ZCTA-month aggregation by multiplying the proportion of either B.1.1.7, B.1.526, or “Other” variants in our sample by the total number of COVID-19 cases.

Maximum spatial and temporal cluster size parameters were set a priori for 10% of the population at risk (24) and one month, respectively. Space-time cluster detection in SaTScan has a noted limitation where the size of clusters cannot change over time (32,33). Given that our data is aggregated to the temporal unit of months (December 2020 – April 2021), setting the maximum temporal cluster size parameter to one month allows clusters to change their shape from month to month by being designated as “new” clusters. A further description of settings for the retrospective multinomial space-time scan statistic can be found in the supplemental materials.

Inverse-distance weighted interpolation and spatial average of SARS-CoV-2 genome sequencing We employed inverse-distance weighted (IDW) interpolation to better visualize how the proportion of COVID-19 cases attributable to each SARS-CoV-2 variant spatiotemporally varies in NY (34). The percentage of COVID-19 cases attributable to each variant in a ZCTA was assigned each ZCTA’s corresponding centroid for the IDW calculation. IDW interpolation generated a continuous surface of values representing the percentage of total COVID-19 cases attributed to B.1.1.7 and B.1.526, which were then averaged over each ZCTA geometry.

The estimated percentage of each SARS-CoV-2 variant generated from IDW interpolation was then multiplied by the total number of COVID-19 cases for each ZCTA and month of our study period to estimate the total number of COVID-19 cases attributable to each variant. Estimated numbers of variant cases were then used to generate geographic mean centers for each month of the study period for visualization (35). Methods for geographic mean center calculation can be found in the supplemental materials.

### Phylogeographic analyses

All B.1.526 genomes from the United States (US) and associated metadata (excluding NY sequences) were downloaded from GISAID (GISAID.org) and randomly subsampled to approximately equal depth as the heaviest sampled NY region in our dataset, with the number of genomes from each state sampled proportionally to their overall frequency in the US. The final dataset included B.1.526 genomes from MA, NJ, PA, CT, CA, FL, MD, MI, MN, and NC, aggregated as “Domestic”, genomes from the five boroughs of NYC (Bronx, Brooklyn, Queens, Staten Island, Manhattan), Long Island, and the Hudson Valley and those from Western NY, the Finger Lakes, the Capital District, and Central NY regions aggregated as “Upstate”. Genomes were aligned in mafft v7.475 (36) with problematic sites masked according to (https://github.com/W-L/ProblematicSites_SARS-CoV-2). A maximum likelihood phylogeny was generated in IQTree v1.6.12 (37) with 1000 ultrafast bootstrap replicates (38) and timecalibrated in TreeTime v0.7.6 (39). This tree served as the fixed tree for ancestral state reconstruction in Beast v2.6.2 (40) to infer timing and source of B.1.526 introductions within the state of NY. The Bayesian analysis was allowed to run for > 4 million generations and monitored in Tracer until the effective sample size of all parameters >= 200 and the MCMC chain appeared to reach stationarity.

A B.1.1.7 phylogeographic analysis was conducted in the same manner with the states inferred for a fixed topology over 6 million generations in BEAST2 under an exponential coalescent model until all ESS reach >= 200. The final dataset included B.1.1.7 genomes from MA, PA, CT, NJ, CA, and FL grouped together as “Domestic”, the NYC, Long Island, MidHudson and Finger Lakes regions of NY, “Southwest NY” composed of the Southern Tier and Western regions of NY, and “Northern NY” comprised of the Capital District, Mohawk Valley,

### Central NY, and the North Country

Maximum clade credibility trees for B.1.526 and B.1.1.7 were generated in TreeAnnotator v.2.6.2 (40) with a 10% burn-in. The number of introductions between locations was summarized by Baltic (https://github.com/evogytis/baltic) by adopting the exploded tree script for Python 3. Only introductions with a posterior probability of 0.7 >= were considered. Trees were visualized in FigTree v1.5.5 (http://tree.bio.ed.ac.uk/software/figtree/) and ggtree (41) for R v4.1.0 (http://www.R-project.org) (see supplementary information for additional details).

## Data Availability

The sequence data that support the findings of this study are openly available in GISAID.org. The disease incidence and location data are not publicly available due to their containing information that could compromise the privacy of patients.

http://www.GISAID.org

## IRB Approval

This work was approved by the New York State Department of Health Institutional Review Board, under study numbers 02-054 and 07-022.

## Acknowledgements

The authors gratefully acknowledge the Advanced Genomic Technologies Core of the Wadsworth Center where all next generation sequencing was performed. We also graciously thank the New York State clinical laboratories that submitted SARS-CoV-2 positive specimens to Wadsworth for sequence analysis, all originating and submitting laboratories for their SARS-CoV-2 sequence contributions to the GISAID database, Wadsworth Center’s Virology Laboratory for initial processing of specimens and the Bioinformatics Core for sequence processing and analysis.

