## Supplemental methods and data for "Spatiotemporal analyses illuminate the competitive advantage of a SARS-CoV-2 variant of concern over a variant of interest"

**Supplemental materials:**

*Geographic mean center calculation*

The geographic mean center of total cases and estimated variant cases of COVID-19 were calculated using equation 1.

Eq. 1: $\frac{\sum_i^N x_i z_i}{\sum_i^N z_i}, \frac{\sum_i^N y_i z_i}{\sum_i^N z_i}$

Where $x_i$ and $y_i$ denote latitude and longitude values of a ZCTA centroid, respectively, and zi denotes the number of cases recorded or estimated for a ZCTA. Centroid calculation, spatial averaging, IDW methods and maps were performed using the 'sf', 'raster', 'gstat', and 'tmap' packages in RStudio version 4.0.2, respectively (Hijmans, 2020; Pebesma, 2004, 2018; RStudio Team, 2020; Tennekes, 2018)

*Retrospective multinomial space-time scan statistic*

The procedure for the multinomial scan statistic implemented in SaTScan from Jung et al. (2010) is described below, using terms that apply to our research questions. The multinomial scan statistic assesses the null hypothesis of no clustering by globally testing whether the probability of acquiring a specific variant of SARS-CoV-2 relative to all variants of SARS-CoV-2 is the same in all parts of the study area. The rejection of the global null hypothesis permits for the scanning of a specific region and regions while testing the same null hypothesis locally. Specifically, the space-time scan procedure operates by searching for clusters in a "moving cylinder" fashion, such that the base of the cylinder is the spatial scan, while the height of the cylinder indicates the temporal scan. As the cylinder moves throughout the spatiotemporal study region, the test statistic is calculated for each scanning window, and the window that maximizes the likelihood ratio test statistic is selected as the most likely cluster. For specific details on the likelihood function and test statistic, see Jung et al., 2010 .

The moving cylinder method employed by SaTScan presents a key limitation for use examining disease outbreaks. The geometry of a cylinder does not allow for the change in the spatial extent of a cluster throughout time, as would be expected for a disease cluster that is spreading (Takahashi et al., 2008). Methodologies have been proposed to alleviate this problem, including the "square pyramid" method and the "flexible space-time scan statistic" (Iyengar, 2005; Takahashi et al., 2008). Neither the square pyramid nor the flexible space-time scan statistics were available in the SaTScan software, thus, we elected to reduce our maximum temporal cluster size to be equivalent to our time precision. Additionally, adjusting the population at risk parameter when using the multinomial scan statistic sets an upper bound for the size of a cluster according to the number of cases it will include, rather than the population at risk. In this way, clusters resulting from our analysis will not include more than 10% of the total cases during our specific time aggregation units of one month.

*Illumina library preparation and sequencing:*

Extracted RNA was processed for whole genome sequencing with a modified ARTIC protocol (artic.network/ncov-2019) in the Applied Genomics Technology Core at the Wadsworth Center. Briefly, cDNA was synthesized with SuperScript™ IV reverse transcriptase (Invitrogen, Carlsbad, CA, USA) and random hexamers. Amplicons were generated by pooled PCR with two premixed ARTIC V3 primer tools (Integrated DNA Technologies, Coralville, IA, USA).

Additional primers to supplement those showing poor amplification efficiency (github.com/artic-network/artic-ncov2019/tree/master/primer_schemes/nCoV2019) were added separately to the pooled stocks. PCR conditions were 98°C for 30 seconds, 24 cycles of 98°C for 15 seconds/63°C for 5 minutes, and a final 65°C extension for 5 minutes. Amplicons from pool 1 and pool 2 reactions were combined and purified by AMPure XP beads (Beckman Coulter, Brea, CA, USA) with a 1X bead-to-sample ratio and eluted in 10mM Tris-HCl (pH 8.0). The amplicons were quantified using Quant-IT™ dsDNA Assay Kit on an ARVO™ X3 Multimode Plate Reader (Perkin Elmer, Waltham, MA, USA). Illumina sequencing libraries were generated using the Nextera DNA Flex Library Prep Kit with Illumina Index Adaptors and sequencing on a MiSeq instrument (Illumina, San Diego, CA, USA).

Bioinformatics processing

Illumina libraries were processed with ARTIC nextflow pipelines (github.com/connor-lab/ncov2019/articnf/tree/illumine, last updated April 2020) as previusly described (Alpert et al., 2021). Reads were trimmed with TrimGalore (github.com/FelixKrueger/TrimGalore) and aligned to the reference assembly MN908947.3 (Wuhan-1) by BWA (Li & Durbin, 2010). Primers were trimmed with iVar (Grubaugh et al., 2018) and variants were called with samtools mpileup function (Li et al., 2009), the output of which was used by iVar to generate consensus sequences. Positions were required to be covered by a minimum depth of 50 reads and variants were required to be present at a frequency $\geq$0.75.

Lineages were determined by GISAID using Pangolin software 29, last updated May 27, 2021 (Rambaut et al., 2020). At the time of this analysis, B.1.526 was divided into a B.1.526 parent lineage and sublineages B.1.526.1, B.1.526.2, and B.1.526.3, which we analyzed separately in the multinomial scan analysis. Pangolin has since collapsed the sublineages and reassigned all to B.1.526.

### Phylogeographic analyses

All B.1.526 genomes from the United States (US) and associated metadata (excluding NY sequences) were downloaded from GISAID (GISAID.org) and randomly subsampled to approximately equal depth as the heaviest sampled NY region in our dataset, with the number of genomes from each state sampled proportionally to their overall frequency in the US. Genomes were aligned in mafft v7.475 (Katoh & Standley, 2013) with problematic sites masked according to (https://github.com/W-L/ProblematicSites_SARS-CoV2). Putative transmission clusters were identified by TreeCluster v1.0.3 (Balaban et al., 2019) with a threshold free approach and only one representative genome was selected from each cluster if 1) all genomes derived from the same state within a one week time period or 2) all genomes derived from the same NY county within a one week time period to reduce the size of the dataset. After generating an initial ML tree in IQTree v1.6.12 (Nguyen et al., 2015) under a GTR+G substitution model, it became apparent that most states contributed minimally or not at all to the number of B.1.526 introductions into NY. It also appeared that most B.1.526 viral circulation occurred between NY and geographically proximal states (Petrone et al., 2021). As the focus of our paper was mainly to document the spread of B.1.526 within NY as compared to B.1.1.7, we further reduced our dataset to include only states with the greatest number of sequenced B.1.526 cases and neighboring states to NY. Temporal signal was confirmed by TempEst v1.5.3 (Rambaut et al., 2016) and genomes with residuals > 0.005 were removed. The final dataset included B.1.526 genomes from MA, NJ, PA, CT, CA, FL, MD, MI, MN, and NC, aggregated as “Domestic”. Because B.1.526 likely originated within the Metro region (as defined in Figure 1B), we elected to keep the five boroughs of NYC (Bronx, Brooklyn, Queens, Staten Island, Manhattan) as well as Long Island and Hudson Valley as distinct to infer the geographic origin of B.1.526 and determine transmission dynamics in this epicenter. The other regions of NY had either no or a considerably lower number of sequenced cases of B.1.526, which is consistent with the incidence of the variant in those regions. Thus, Western NY, the Finger Lakes, the Capital District, and Central NY regions were aggregated as “Upstate”. A second ML tree was generated for this reduced dataset in IQTree with 1000 ultrafast bootstrap replicates (Minh et al., 2013). This tree was then input into TreeTime v0.7.6 (Sagulenko et al., 2018) to estimate a molecular clock and re-root the tree with the least-squares method. The time-calibrated tree was input as the fixed tree for discrete ancestral state reconstruction in BEAST2 v2.6.2 (a method previously validated by Alpert et al., 2021; Bouckaert et al., 2019; Lemey et al., 2009). The phylogeographic analysis ran under a GTR+ G4 substitution model with the molecular clock set to 4.0E-04 substitutions per site per year and an exponential coalescent population model. The Bayesian analysis was allowed to run for > 4 million generations and monitored in Tracer until the effective sample size of all parameters >= 200 and the MCMC chain appeared to reach stationarity.

A B.1.1.7 phylogeographic analysis was conducted in the same manner with the following exceptions: the tree was initially rooted with a P.1 (Gamma) representative as B.1.1.7 cases in NY had multiple origins, the five boroughs of NYC were included as the same region as it has been established that B.1.1.7 was introduced several times from non-NYC locations, the Capital District, Mohawk Valley, Central NY, and the North Country were aggregated as “Northern NY” given their proximity to each other, Western NY and its neighboring region, the Southern Tier, were grouped together as “Southwestern NY”, the Finger Lakes, the Hudson Valley, and Long Island remained distinct. B.1.1.7 locations required different coding than

B.1.526 due to the substantial differences in sample sizes. For example, genomes from the Finger Lakes accounted for over 25% of the B.1.1.7 data but less than 2% of the data for B.1.526 from NY. MA, PA, CT, NJ, CA, and FL were grouped together as “Domestic” sources of B.1.1.7. Ancestral states were inferred for a fixed topology over 6 million generations in BEAST2 under an exponential coalescent model until all ESS reach >= 200. Maximum clade credibility trees for B.1.526 and B.1.1.7 were generated in TreeAnnotator v.2.6.2 (Bouckaert et al., 2019) with a 10% burn-in. The number of introductions between locations was summarized by Baltic (https://github.com/evogytis/baltic) by adopting the exploded tree script for Python 3. Only introductions with a posterior probability of 0.7 >= were considered. Trees were visualized in FigTree v1.4.4 (http://tree.bio.ed.ac.uk/software/figtree/) and ggtree (Yu et al., 2017) for R v4.1.0 (http://www.R-project.org).

Supplementary figures and tables

Figure 1:

A) Proportion of B.1.1.7, B.1.526 and other lineages by New York State region by week, 12/1/2020 – 4/26/2021
B) NYSDOH Regions of NY
C) Number of specimens and percent of cases sequenced by week, 12/1/2020 – 4/26/2021.

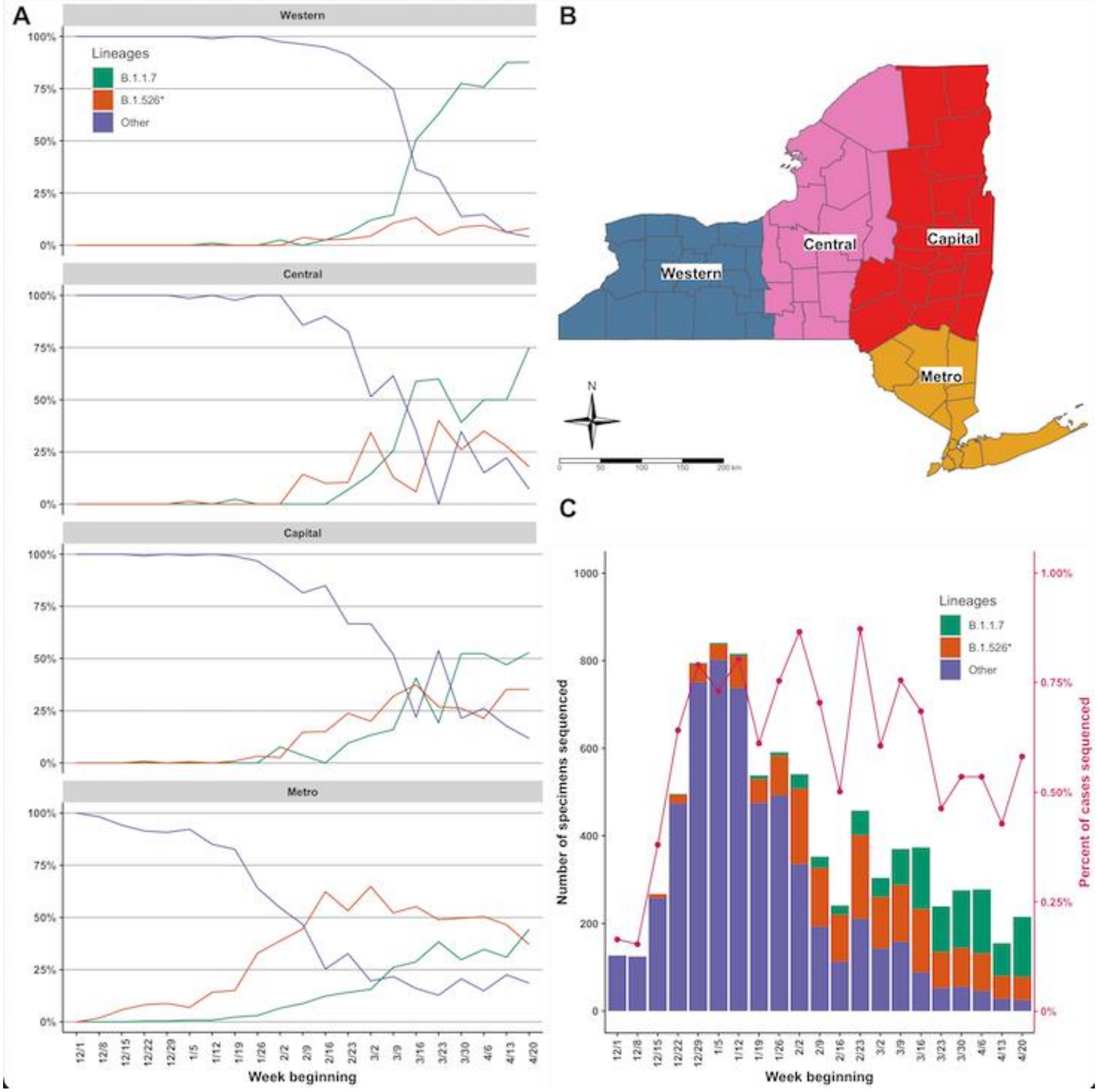

Table 1: Multinomial cluster analysis cluster-specific relative risks

| Cluster | Month | Lineage | | | | | |
|---|---|---|---|---|---|---|---|
| | | B.1.1.7 | B.1.526 | B.1.526.1 | B.1.526.2 | B.1.526.3 | Other |
| **1** | December | 0 | 0 | 0 | 0 | 0 | **1.56** |
| **2** | December | 0 | 0 | 0 | 0 | 0 | **1.56** |
| **3** | March | **2.83** | **4.11** | 0.36 | **1.48** | 0 | 0.13 |
| **4** | March | **1.32** | **2.82** | **4.44** | **3.77** | 0 | 0.18 |
| **5** | March | **4.59** | **1.66** | **1.12** | **1.71** | 0 | 0.19 |
| **6** | April | **7.49** | 0.29 | 0.27 | 0.54 | 0 | 0.11 |

Relative risk (RR) greater than 1.00 is bold.

Table 2:

**Table 2.** Number of B.1.526 introductions from various New York and Domestic locations. The number of introductions that occurred to a location (To) from a source (From) at >= 0.7 posterior probability for ancestral location. Percentage of Total, the proportion of the total number of introductions for all locations; Region Totals, the sample size of each location and the total proportion of introductions contributed from this area.

| From | To | Introductions | Percentage of Total | Region Totals |
|---|---|---|---|---|
| Bronx | Domestic | 50 | 19.7 | 222/63.8 |
| Bronx | Hudson | 24 | 9.4 | |
| Bronx | Kings | 11 | 4.3 | |
| Bronx | Long Island | 17 | 6.7 | |
| Bronx | Manhattan | 18 | 7.1 | |
| Bronx | Queens | 19 | 7.5 | |
| Bronx | Staten Island | 2 | 0.8 | |
| Bronx | Upstate | 21 | 8.3 | |
| Domestic | Bronx | 3 | 1.2 | 290/6.7 |
| Domestic | Hudson | 8 | 3.1 | |
| Domestic | Kings | 0 | 0.0 | |
| Domestic | Long Island | 4 | 1.6 | |
| Domestic | Manhattan | 0 | 0.0 | |
| Domestic | Queens | 0 | 0.0 | |
| Domestic | Staten Island | 0 | 0.0 | |
| Domestic | Upstate | 2 | 0.8 | |
| Hudson | Bronx | 5 | 2.0 | 128/9.4 |
| Hudson | Domestic | 15 | 5.9 | |
| Hudson | Kings | 0 | 0.0 | |
| Hudson | Long Island | 2 | 0.8 | |
| Hudson | Manhattan | 0 | 0.0 | |
| Hudson | Queens | 0 | 0.0 | |
| Hudson | Staten Island | 1 | 0.4 | |
| Hudson | Upstate | 1 | 0.4 | |
| Brooklyn | Bronx | 2 | 0.8 | 39/2.4 |
| Brooklyn | Domestic | 2 | 0.8 | |
| Brooklyn | Hudson | 0 | 0.0 | |
| Brooklyn | Long Island | 0 | 0.0 | |
| Brooklyn | Manhattan | 1 | 0.4 | |
| Brooklyn | Queens | 1 | 0.4 | |
| Brooklyn | Staten Island | 0 | 0.0 | |
| Brooklyn | Upstate | 0 | 0.0 | |
| Long Island | Bronx | 0 | 0.0 | 78/2.8 |
| Long Island | Domestic | 5 | 2.0 | |
| Long Island | Hudson | 0 | 0.0 | |
| Long Island | Kings | 0 | 0.0 | |
| Long Island | Manhattan | 1 | 0.4 | |
| Long Island | Queens | 0 | 0.0 | |
| Long Island | Staten Island | 1 | 0.4 | |
| Long Island | Upstate | 0 | 0.0 | |
| Manhattan | Bronx | 5 | 2.0 | 49/4.7 |
| Manhattan | Domestic | 4 | 1.6 | |
| Manhattan | Hudson | 0 | 0.0 | |
| Manhattan | Kings | 2 | 0.8 | |
| Manhattan | Long Island | 0 | 0.0 | |
| Manhattan | Queens | 1 | 0.4 | |
| Manhattan | Staten Island | 0 | 0.0 | |
| Manhattan | Upstate | 0 | 0.0 | |
| Queens | Bronx | 2 | 0.8 | 81/6.3 |
| Queens | Domestic | 5 | 2.0 | |
| Queens | Hudson | 0 | 0.0 | |
| Queens | Kings | 4 | 1.6 | |
| Queens | Long Island | 1 | 0.4 | |
| Queens | Manhattan | 3 | 1.2 | |
| Queens | Staten Island | 0 | 0.0 | |
| Queens | Upstate | 1 | 0.4 | |
| Staten Island | Bronx | 0 | 0.0 | 12/0.8 |
| Staten Island | Domestic | 2 | 0.8 | |
| Staten Island | Hudson | 0 | 0.0 | |
| Staten Island | Kings | 0 | 0.0 | |
| Staten Island | Long Island | 0 | 0.0 | |
| Staten Island | Manhattan | 0 | 0.0 | |
| Staten Island | Queens | 0 | 0.0 | |
| Staten Island | Upstate | 0 | 0.0 | |
| Upstate | Bronx | 3 | 1.2 | 81/3.1 |
| Upstate | Domestic | 2 | 0.8 | |
| Upstate | Hudson | 1 | 0.4 | |
| Upstate | Kings | 0 | 0.0 | |
| Upstate | Long Island | 1 | 0.4 | |
| Upstate | Manhattan | 0 | 0.0 | |
| Upstate | Queens | 1 | 0.4 | |
| Upstate | Staten Island | 0 | 0.0 | |

Table 3:

**Table 3.** Number of B.1.1.7 introductions from various New York and Domestic locations. The number of introductions that occurred to a location (To) from a source (From) at >= 0.7 posterior probability for ancestral location. Percentage of Total, the proportion of the total number of introductions for all locations; Region Totals, the sample size of each location and the total proportion of introductions contributed from this area.

| From | To | Introductions | Percentage of Total | Region Totals |
|---|---|---|---|---|
| Northern | Domestic | 6 | 2.4 | 149/6.3 |
| Northern | Finger Laeks | 4 | 1.6 | |
| Northern | Hudson | 3 | 1.2 | |
| Northern | Long Island | 1 | 0.4 | |
| Northern | NYC | 2 | 0.8 | |
| Northern | SouthWest | 0 | 0.0 | |
| Domestic | Northern | 26 | 10.2 | 362/38.8 |
| Domestic | Finger Laeks | 30 | 11.8 | |
| Domestic | Hudson | 9 | 3.5 | |
| Domestic | Long Island | 5 | 2.0 | |
| Domestic | NYC | 15 | 5.9 | |
| Domestic | SouthWest | 14 | 5.5 | |
| Finger Lakes | Northern | 2 | 0.8 | 239/7.1 |
| Finger Lakes | Domestic | 7 | 2.7 | |
| Finger Lakes | Hudson | 0 | 0.0 | |
| Finger Lakes | Long Island | 4 | 1.6 | |
| Finger Lakes | NYC | 1 | 0.4 | |
| Finger Lakes | SouthWest | 4 | 1.6 | |
| Hudson | Northern | 1 | 0.4 | 78/14.5 |
| Hudson | Domestic | 5 | 2.0 | |
| Hudson | Finger Laeks | 0 | 0.0 | |
| Hudson | Long Island | 7 | 2.7 | |
| Hudson | NYC | 24 | 9.4 | |
| Hudson | SouthWest | 0 | 0.0 | |
| Long Island | Northern | 3 | 1.2 | 130/10.2 |
| Long Island | Domestic | 8 | 3.1 | |
| Long Island | Finger Laeks | 1 | 0.4 | |
| Long Island | Hudson | 1 | 0.4 | |
| Long Island | NYC | 11 | 4.3 | |
| Long Island | SouthWest | 2 | 0.8 | |
| NYC | Northern | 4 | 1.6 | 181/21.6 |
| NYC | Domestic | 26 | 10.2 | |
| NYC | Finger Laeks | 2 | 0.8 | |
| NYC | Hudson | 10 | 3.9 | |
| NYC | Long Island | 12 | 4.7 | |
| NYC | SouthWest | 1 | 0.4 | |
| SouthWest | Northern | 0 | 0.0 | 56/1.6 |
| SouthWest | Domestic | 1 | 0.4 | |
| SouthWest | Finger Laeks | 3 | 1.2 | |
| SouthWest | Hudson | 0 | 0.0 | |
| SouthWest | Long Island | 0 | 0.0 | |
| SouthWest | NYC | 0 | 0.0 | |
